# Assessing the risk stratification of breast cancer polygenic risk scores in two Brazilian samples

**DOI:** 10.1101/2022.09.09.22279721

**Authors:** R.A.S Barreiro, TF Almeida, CS Gomes, F Monfardini, AA Farias, GC Tunes, GM Souza, E Duim, JS Correia, AVC Coelho, MP Caraciolo, YAO Duarte, M Zatz, E Amaro, JB Oliveira, BD Bitarello, H Brentani, MS Naslavsky

## Abstract

Polygenic risk scores (PRS) for breast cancer (BC) have a clear clinical utility in risk prediction. PRS transferability across populations and ancestry groups is hampered by population-specific factors, ultimately leading to differences in variant effects, such as linkage disequilibrium (LD) and differences in variant frequency (AF-diff). Thus, locally-sourced population-based phenotypic and genomic datasets are essential to assess the validity of PRS derived from signals detected across populations. Here, assess the transferability of a BC PRS composed of 313 risk variants (313-PRS) in two Brazilian tri-hybrid admixed ancestries (European, African and Native American) whole-genome sequenced cohorts. We computed 313-PRS in both cohorts (n=753 and n=853) versus the UK Biobank (UKBB, n=264,307) as reference. We show that although the Brazilian cohorts have a high European (EA) component, with AF-diff and to a lesser extent LD patterns like those found in EA populations, the 313-PRS distribution is inflated when compared to that of the UKBB, leading to potential overestimation of PRS-based risk if EA is taken as a standard. Interestingly, we find that case-controls lead to equivalent predictive power when compared to UKBB-EA samples with AUROC values of 0.66-0.62 compared to 0.63 for UKBB.

## INTRODUCTION

Assessing polygenic risk scores (PRS) transferability across ancestries is crucial if genetic risk stratification at the clinical level is to be implemented in populations with ancestry profiles that differ from those assessed in genome-wide association studies (GWAS) ^1^. PRS derived by using individuals from a given genetic ancestry yield reduced predictions when tested in individuals of other ancestries, including those with admixed genomes, due to a complex interaction between differences in linkage disequilibrium (LD) and allele frequencies (AF) among populations, as well as population specific gene-environment and epistatic effects - all of which contribute to each disease or trait’s genetic architecture. The consequences of different populations having these population-specific parameters ultimately lead to differences in variant effects, and overall PRS performance. To address this issue, novel statistical genetic techniques are being applied ^2,3^ and diverse new genetic databases are becoming available ^4,5^. Moreover, improvements to this performance discrepancy outside of the tested ancestry have been proposed ^6^ and a few studies have focused on admixed cohorts ^3,7^, but none provided a definitive solution.

The potential use of PRS for improvements in disease prevention and management has been a prolific area of scientific investigation in the past decade, enabled by the availability of large datasets such as the UK BioBank (UKBB). While an essential resource, the UKBB and other major Biobanks are heavily biased in their ancestry representation ^1,3,8^. Thus, much of the aforementioned assessments of transferability of PRS across ancestries rely on smaller cohorts that contain a majority of underrepresented ancestry groups such as Latino/Latinx/Hispanic, African/Black/African-American, Native American, and East Asian.

As cohorts of over 1,000 admixed individuals with comprehensive phenotypic characterization and high-coverage whole–genome sequencing (WGS) data arise in Brazil ^4,5^, evaluating the distribution of PRS derived from large European GWAS becomes an important first step prior to clinical implementation. Breast cancer (BC) is the worldwide leading cause of death in women and a complex disorder of multifactorial inheritance. While up to 10% of BC cases are attributed to large effect pathogenic variants segregating in families, most affected individuals are not carriers, suggesting that sporadic occurrence is influenced by a polygenic combination of small to moderate effect variants ^1^ with familial aggregation and heritability reaching 55% ^9^.

For BC, PRS derived from European ancestry (EA) individuals do have predictive ability even in African ancestry (AA) populations, but performance dropped significantly compared to the EA population ^10^. A 313-SNP BC PRS (“313-PRS”) trained in EA populations was shown to have improved predictive power compared to previously described breast cancer PRS ^11^. Moreover, the 313-PRS has greater discriminatory power compared to risk prediction models based only on classical risk factors in EA populations and its incorporation in BC predictive models provide a greater level of risk stratification in the general population ^12^. Thus, “risk-stratified” breast cancer screening could potentially contribute to breast cancer early detection and the 313-PRS have been proposed as a useful tool to improve screening efficiency ^13^.

Here, we investigate the potential for the 313-PRS ^11^ as a screening tool for BC in two Brazilian admixed cohorts. We compare the PRS distributions between these Brazilian cohorts and the UKBB (composed largely of EA individuals) and evaluate the predictive power of a model including 313-PRS as predictor. We find that even though the Brazilian cohorts have a relatively high EA component mirrorer by both LD and AF-diff, the overall PRS distribution was inflated when compared to the UKBB, leading to potential overestimation of PRS-based risk if these cohorts are assumed to have similar PRS distributions as the UKBB. Nevertheless, stratification by outcomes led to an equivalent area under the receiver operator curve (AUROC) values when compared to UKBB. We discuss the limitations and implications of these findings below.

## MATERIALS & METHODS

### Genotypic and phenotypic data

Data for the present study comes from female samples of two Brazilian cohorts: 1) The Health, Well-being and Aging Study (SABE, acronym in Portuguese), a census-based sample of individuals aged above 60 years old living in São Paulo, Brazil, with 21 cases of BC and 732 controls^5^; 2) The Rare Genomes Project (GRAR)^4^, a nationwide sample of probands ascertained by being at risk for Mendelian disorders recruited after clinical evaluation, comprising 322 cases of BC and 531 controls. To address potential confounders in GRAR, we removed individuals with related cancers such as ovarian and pancreatic cancer and BC-cases with causative monogenic variants. We also used the UKBB imputed dataset ^14^ (Project 74348), with 11,245 cases and 253,062 controls for comparison. See **Supplementary Methods** for details.

### Data processing

We mapped SABE and GRAR WGS data to the human reference genome hg38 with DRAGEN-GATK (Illumina, version 3.6.4 or superior) and genotyped using Illumina’s DRAGEN DNA pipeline as previously described ^4,5^. Of 313 BC-related SNPs, 308 were present in all cohorts and used for downstream analyses. We call this set 313-SNPs for ease of reading.

### Ancestry, LD, PRS, and modeling

We inferred global ancestry for all cohorts using Principal Component Analysis (PCA) and supervised ADMIXTURE ^15^. We used 1000 Genomes Project Phase 3 (1KGP3) individuals classified as African (AFR), European (EUR) and East Asian (EAS) for ancestry-specific PRS distributions comparisons (see **Supplementary Methods**).

We collected overlapping variants in all sets of individuals within a region of 500 kbp upstream and downstream of the 313-PRS and used varLD^16^ to perform LD comparisons between SABE and GRAR, as well as with 1KGP3 individuals grouped according to the three main continental ancestries (AFR, EUR, EAS).

We calculated the 313-PRS separately for all cohorts and 1000KGP3 populations, including both unaffected and affected women from SABE, GRAR, and the UKBB. Each individual’s PRS was calculated as the sum of the risk alleles dosage, *G*, times its effect-size (*w*) for every variant (*i*) (PRS = ∑_*i*_*w*_*i*_*G*_*i*_), using previously inferred weights from an EA-cohort^11^.

To evaluate the PRS regression models performance in the Brazilian and UKBB cohorts we calculated area under receiver operator curve (AUROC), odds-ratio per standard deviation (ORperSD) and Partial-*R*^2^ between models with and without the PRS. Confidence intervals were calculated using 1,000 bootstraps. Scripts to replicate these analyses are available at: https://github.com/Varstation/313prs-bc-grar-sabe

## RESULTS

Since the two Brazilian cohorts were sampled differently, we explored in a comparative way variables that are known to affect PRS transferability across cohorts: global ancestry composition, 313-PRS risk allele frequency (RAF), and LD patterns in genomic regions linked to risk variants.

Patterns of global ancestry inferred both by PCA (Figure 1A) and supervised ADMIXTURE (Supplementary Figure 1) show that GRAR and SABE share similar ancestry patterns, with most individuals distributed as a continuum between AFR and EUR ancestries and a small number of individuals with higher proportions of EAS ancestry. Additionally, both cohorts share similar patterns of genome-wide principal components (Supplementary Figure 2).

**Figure 1.**
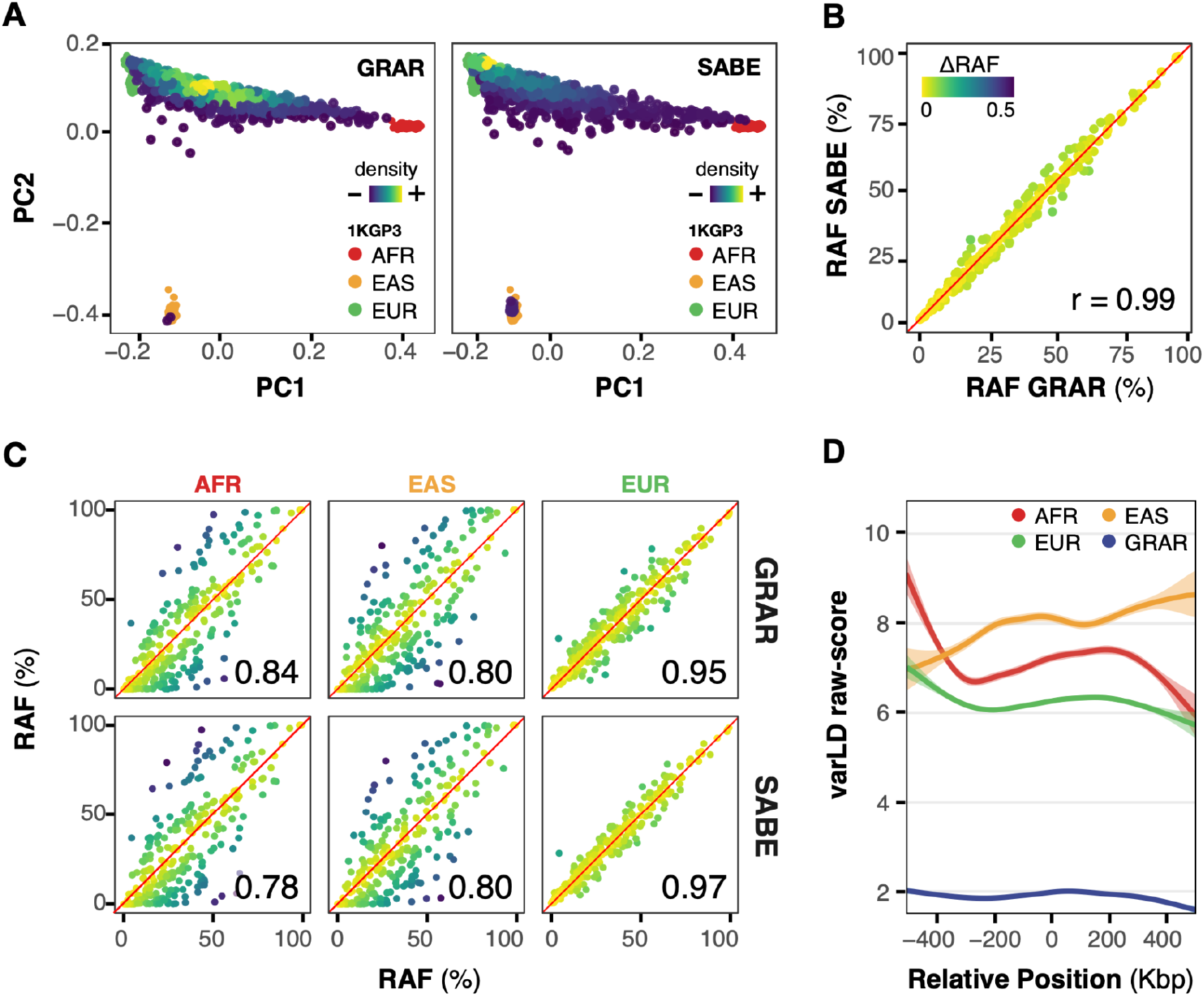
Allelic frequency and LD patterns for two Brazilian cohorts (SABE and GRAR) **(A)** Principal component (PC) analysis of GRAR (left) and SABE (right). Plots show PC1 and PC2 in the x- and y-axis, respectively. Different colors refer to 1KGP3 ancestry-specific subsets of individuals. **(B)** Risk allele frequency (RAF) for GRAR (x-axis) and SABE (y-axis); r, Pearson’s correlation coefficient. **(C)** RAF correlation between Brazilian cohorts and 1KGP3-EUR, -AFR, -EAS (from left to right). GRAR (top row); SABE (bottom row); *r*, (Pearson’s correlation coefficient). 1KGP3. **(D)** Raw varLD scores for 1Mbp windows centered across the 313-PRS BC risk SNPs. Lines show varLD calculated between SABE and 1KGP3-EUR; 1KGP3-AFR; 1KGP3-EAS; GRAR (dark blue).; x-axis, 0 represents the risk SNP coordinate. In panels A, C, D, AFR (red), EUR (green), and EAS (orange) refer to the African, European, and East Asian subsets of individuals from the 1,000 Genomes Phase 3 (1KGP3).

GRAR and SABE have similar RAFs for the BC 313-SNPs (Figure 1B), and both cohorts have an overall higher correlation with RAFs calculated for the EA subset of 1KGP3 individuals (Figure 1C) than with the AFR or EAS ancestry subsets (Pearson’s cor >= 0.95 in both comparisons). Both cohorts share the same correlation with the EUR 1KGP3 subset (cor = 0.80), whereas GRAR has a higher RAF correlation with AFR than SABE (cor=0.84 and cor=0.78, respectively). LD patterns surrounding the 313 risk SNPs are more similar between SABE and GRAR than any of the other pairwise comparisons (Figure 1D).

Prior to PRS analyses, we explored potential effects of different demographic characteristics between the cohorts (Supplementary Table 1). Because the SABE cohort has a small number of cases (n=21), we simulated sampling and case-control balances of SABE with bootstrap analyses on UKBB (Supplementary Figure 3) and did not observe bias regarding the small case numbers present in SABE data.

Next, we calculated 313-PRS for GRAR, SABE, GRAR+SABE (G+S) and UKBB cohorts. We compared the 313-PRS PRS distribution obtained from all individuals in GRAR (n=853), SABE (n=753) and UKBB (n=264,307). The Brazilian cohorts and non-European ancestry 1KGP3 populations – AFR (n=258) and EAS (n=260) – had a distribution of 313-PRS values shifted towards higher values than the UKBB and EUR-1KGP3 (n=263), which had similar distributions (Wilcoxon sum rank test, *p* = 0.69) between them and statistically significant differences from all other cohorts (*p* < 0.0001 for all comparisons (Figure 2A).

**Figure 2.**
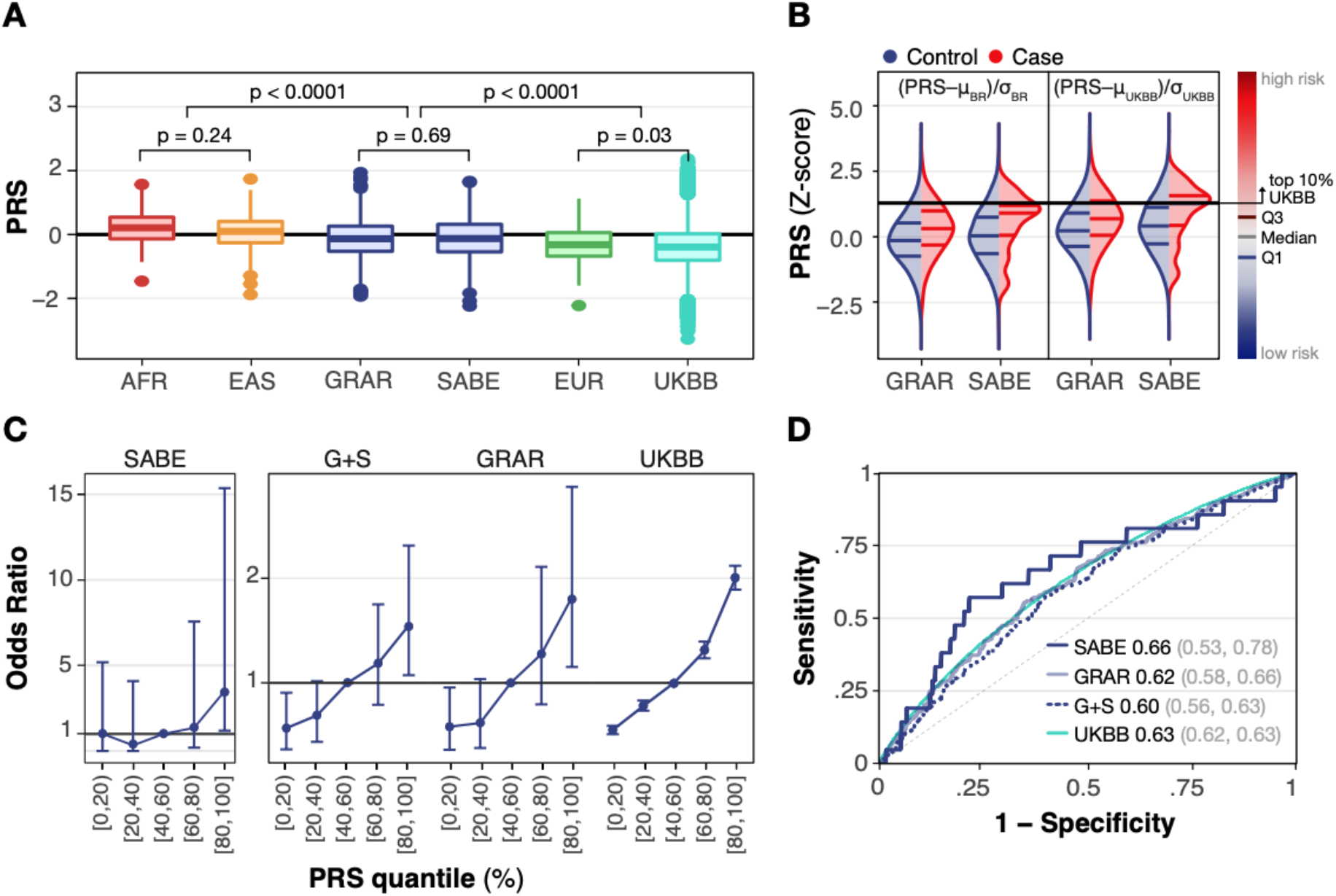
Overall PRS distribution and model assessment. **(A)** 313-PRS distributions for SABE, GRAR, UKBB, and three subsets from the 1KGP3 (AFR, EUR, EAS). **(B)** Normalized 313-PRS based on G+S (on the left) and UKBB (on the right). The black line represents the top 10% UKBB-PRS threshold. **(C)** Odds-ratios based on 313-PRS values for SABE, GRAR, SABE and GRAR combined (G+S), and UKBB. **(D)** Receiver-operator curves (ROC) of the predictive power of a logistic regression model with 313-PRS as the sole predictor of breast cancer. Numbers in the figure indicate the AUROC and values in parenthesis are the 95% confidence intervals.

In both Brazilian cohorts, the mean 313-PRS was significantly higher in cases than controls (Wilcoxon rank sum test, GRAR: *p* < 0.0001, SABE: *p* = 0.012). When we performed a Z-score normalization using the UKBB mean and standard deviation (SD) we found that the third quartiles of SABE and GRAR surpassed the top 10% UKBB PRS (Figure 2B), suggesting that PRS assessment without the specific population mean and sd will inflate the number of individuals inferred to be in higher risk categories.

SABE had a wider range of odds-ratio (OR) values while G+S, GRAR and UKBB share scales of OR values (up to OR∼2) distributed along PRS quantile bins (Figure 2C). In all cohorts, the comparison of OR between quantiles showed that higher PRS quantiles are associated with higher BC OR.

We found similar PRS predictive power for GRAR, G+S, and UKBB cohorts, considering AUROC (only 313-PRS). Whereas SABE showed a higher AUROC (Figure 2D) it has a lower predictive power according the Partial-*R*^2^ metrics (full model including age+5PCs compared to model without 313-PRS) with bootstrap analysis (Supplementary Figure 4). It is important to note that SABE cohort is on average older (72 yrs vs. 40 yrs) and less affected than the GRAR cohort (2.8% cases vs. 38% cases) and the potential effects of these differences could contribute to the results.

## DISCUSSION

We observed that the BC 313-PRS values were on average higher in the two Brazilian cohorts and non-European ancestry groups from the 1KGP3 compared to the UKBB and 1KGP3-EUR. Similar findings have been reported in other BC PRS and specifically for the BC 313-PRS.

One study assessed the transferability of a standardized 313-PRS ^17^ to East Asian ancestry cohorts. The standardization was based on EA values and aimed at enabling comparisons of the PRS performance across populations. However, the authors concluded that a better approach is to standardize the 313-PRS to Asian ancestry values instead, thus minimizing overfitting. We performed a similar approach, by standardizing the 313-PRS to both the EA and Brazilian distributions. We find that, indeed, the former would lead to an inflated number of individuals placed in the high risk groups compared to the latter.

Fritsche and colleagues constructed a BC PRS specifically for the EA subset of UKBB individuals using a sparse set of 334 SNPs (similar to our approach) and a different approach leveraging the weight of 1.1 million SNPs based on population-specific LD patterns for the score and tested both PRSs in UKBB individuals from Asian and African ancestry groups. They observed different distributions in group means of tested BC PRS across European, South Asian, African and East Asian ancestry groups. Their BC-PRS for both PRS approaches were on average higher in non-EUR groups, which we also describe here, but the PRS distribution was nevertheless right-shifted in cases compared to controls and associated with increased continuous ORs when standardized to one SD within each ancestry group ^18^.

Our classification performance was moderate (AUROC=0.60), with an increasing proportion of cases in the top 20% percentile of the 313-PRS distributions in both Brazilian cohorts. A study using the eMERGE network ^19^ to assess the transferability of BC PRS models, including the 313-PRS, compared cohorts with a predominance of EA ancestry to cohorts including women with African and also various proportions of admixed European-African ancestry and self-identified (e.g. not based on genomic ancestry) Latinx women. The AUROCs in self-identified Latinx women for different BC PRS ranged from 0.53 to 0.56 ^19^, compared to 0.63 for the EA cohort. An increasing number of other studies, as well as our own observations, highlight the need to improve the representation of diverse population groups in genomic research cohorts so that population-specific effect sizes can improve PRS prediction power.

Our study has limitations inherent to a general paucity of genomic-level datasets that also include phenotypic data. SABE and GRAR were sampled with different designs and thus they differ in at least two important aspects. The median age of SABE (72 years) is higher than the BC median age of onset (61.8 years) ^20^ and higher than the median age in GRAR (40 years) and the UKBB (58 years). Thus, a survival effect is likely to play a role in differences seen between these cohorts. In fact, unaffected elderly can be considered super-controls, meaning that at a given statistical power, less individuals are required to obtain equivalent effect sizes in case-control setups ^21^. Moreover, the restricted number of cases may affect the results due to low sampling power.

Our findings showed that combining GRAR and SABE led to a loss of predictive power. We interpret this as being due to the important differences highlighted above (age, recruitment approach, family history) and different proportions of cases and controls given that, as shown here, their patterns of LD and RAF are remarkably similar. If large-scale cohorts such as GRAR and SABE are planned and designed in an integrated way, such discrepancies would likely be reduced. On the other hand, our findings also highlight how much the performance metric depends on the sample (cohort), even when ancestry profiles are similar, as highlighted by others.

Our study ultimately provides insights of extreme samples, which may not reflect the expected ancestry-driven reduction in PRS transferability. Given the absence of a valuable large Brazilian population-based biobank, the approach of using available samples with targeted design is an alternative to test PRS transferability. With these important caveats in mind, we note that neither age nor family history (GRAR cases inclusion criterium) are correlated with ancestry components in those cohorts and, as we showed here, the ancestry profiles are remarkably similar.

A reduced PRS predictive power compared to the population or ancestry group from the original is expected due to differences in ancestry composition, allele frequencies and LD patterns across populations. Although both target samples are Brazilian, it is important to consider that the GRAR and SABE cohorts were collected for different purposes and from regions with different demographic histories. Even with different backgrounds both cohorts bear a Brazilian signature being closer to each other than with any other ancestry, suggesting the results could be observed in other Brazilian admixed cohorts. Additional validation on these findings, in particular in correctly ascertaining cases within the highest PRS percentiles, might advocate towards clinical applications on transferred PRS, while large GWAS on each ancestral groups are performed and crucial non-European population-based biobanks are constructed.

## Data Availability

Scripts to replicate these analyses are available at: https://github.com/Varstation/313prs-bc-grar-sabe. SABE genomic data is available at https://ega-archive.org/studies/EGAS00001005052 under reasonable request and after Data Access Agreement is signed by User and User Institution

https://ega-archive.org/studies/EGAS00001005052

## ACKNOWLEDGMENTS AND FUNDING

This research has been conducted using the UK Biobank Resource under Application Number 74348. Funding for Michel Naslavsky, Mayana Zatz and Yeda Duarte at University of São Paulo was provided by grants and fellowships from São Paulo Research Foundation (FAPESP) (CEPID 2013/08028-1, SABE 2014/50649-6, INCT 2014/50931-3), and Conselho Nacional de Desenvolvimento Científico e Tecnológico - CNPq (INCT 465355/2014-5). Funding for Helena Brentani and Catarina S. Gomes at University of São Paulo was provided by grants and fellowships from FAPESP (grant #2020/16376-3 and #2018/18560-6), and the Conselho Nacional de Desenvolvimento Científico e Tecnológico - Brasil (CNPq) grant #310823/2021-8. We thank Varsomics team for support and infrastructure for data processing (https://varsomics.com/). This research was made possible through access to the data and findings generated by the Rare Genomes Project. The Rare Genomes Project is an initiative of Hospital Israelita Albert Einstein in partnership with the Programa de Apoio ao Desenvolvimento Institucional do Sistema Único de Saúde (PROADI-SUS) from the Brazilian Ministry of Health.

Also, we would like to thank the engineering support of Patrícia Campos, Cassio Reginato and Bruno Laget Merino, and the administrative support of Maura Farias.

