## Supplemental material for "Assessing the risk stratification of breast cancer polygenic risk scores in two Brazilian samples"

**METHODS** 1
The two Brazilian cohorts 1
SABE 2
GRAR 2
Genotyping and quality control 3
Ancestry inference 3
Linkage Disequilibrium (LD) 3
Regression model evaluation 4
Model comparisons and potential pitfalls 4
**Supplementary Tables** 5
**Supplementary Figures** 5

### METHODS

#### The two Brazilian cohorts

Data for the present study was derived from female samples of two Brazilian cohorts: SABE and GRAR. All individuals enrolled in SABE and GRAR signed consent forms approved by internal review boards of their corresponding home research institutions.

### SABE

The Health, Well-being and Aging Study (SABE, acronym in Portuguese) cohort was derived from an epidemiological approach including a census-based sample designed by the Pan-American Health Organization (at baseline) to evaluate health and life conditions of the older population in seven countries of Latin America and the Caribbean (Albala, 2005).

SABE followed a longitudinal approach and probabilistic sampling (census-based) of elderly aged 60 and older from the city of São Paulo, with a re-collection every five years including a new cohort with participants between 60 and 65 years of age. The initial sample was obtained in 2000, using a multi-stage design to select 2,143 participants (Cohort A). The primary and secondary sampling units were the city's census tracts and the households, respectively. In 2006, 1,115 individuals were still in the study (second wave), and the new sample added 298 60-65 years old adults to the study (Cohort B). The third wave (2010) included 746 individuals from cohort A, 240 from Cohort B, and 349 new participants 60-65 years old (Lebrão, 2018). DNA from peripheral blood was extracted for all individuals included in the collection of 2010, therefore representing different time-points depending on the cohort (Naslavsky, 2022).

The information queried included sociodemographic characteristics, health and functional status (such as such as frailty, dexterity, balance, and mobility) and chronic diseases such as hypertension, diabetes, chronic pulmonary disorders, heart conditions, brain strokes, joint conditions, osteoporosis, anemia, depression, and cancer.

The inclusion criterion for this study was having a biological sample withdrawn for genetic analysis. A total of 1,200 individuals collected in 2010 were whole-genome sequenced at 30X, resulting in a variant dataset of 1,171 unrelated individuals (Naslavsky, 2022).

### GRAR

The Rare Genomes Project (GRAR) is a program that offers molecular investigation for rare diseases through whole genome sequencing from 16 Rare Disease centers throughout the country. The individuals represented here are those with suspected hereditary cancer syndromes who had a diagnosis of breast cancer (322 cases), and the 531 female individuals (controls) who did not have a diagnosis of breast cancer (BC). All samples were collected from 2020 to 2022. The inclusion criteria for hereditary cancer syndromes were BC before 45 years old, two or more primary tumors at any age, or more than one family member with a related cancer diagnosis. Most cancer cases came from two Northeastern Brazilian states in the Northeast region (Pernambuco and Bahia). DNA was extracted from peripheral blood samples from both cohorts and submitted to whole-genome sequencing (WGS) at 30X in Illumina's sequencing platform.

### Ethics

All participants signed written consent forms. SABE cohort phenotypic and sequencing protocols were approved in several instances by the University of São Paulo Public Health School ethical committee. SABE and UKBB PRS studies were approved by Albert Einstein Israelite Hospital ethical committee (37924920.9.0000.0071)

### Genotyping and quality control

**SABE & GRAR.** For this study, we reprocessed original BAM files {Coelho, 2022} using the same pipeline as GRAR. We mapped SABE and GRAR samples to the human reference genome hg38 with DRAGEN-GATK and genotyped using Illumina's DRAGEN DNA pipeline as previously described [22,23]. We filtered out variants with call rate lower than 0.90, allelic frequency outside the 0.01-0.999 frequency range or significantly deviated from Hardy-Weinberg equilibrium expectations (two-sided Fisher exact test: *p-value* $< 1 \times 10^{-6}$). We also filtered out samples with call rate less than 0.90, resulting in 1,180 SABE individuals, among which 753 are female (21 are positive breast cancer cases and the other 732 served as controls). Since being at risk of Mendelian disease is one of the recruitment criteria for GRAR, further sample filters were performed in this corhort to remove potential pathogenic variant carriers. Participants carrying any SNVs annotated in ClinVar as pathogenic with SNOMED code 254843006 (*Familial cancer of breast (disorder)*) and patients with any other monogenic mutations identified by GRAR project geneticists (as described in Coelho, 2022) were removed, resulting in 322 cases.

**UKBB.** The UKBB microarray imputed dataset was obtained from the UKBB provided *gfetch* tool. We used the same filters described for SABE and GRAR, but also filtered out variants with INFO score below 0.80. The final set of individuals included 264,307 female individuals — 253,062 breast cancer negative (controls) and 11,245 breast cancer positive (cases).

### Ancestry inference

Global ancestry for SABE and GRAR was inferred using principal component analysis (PCA) with Hail *hwe_normalized_pca()* function (https://hail.is/; https://github.com/hail-is/hail), and supervised ADMIXTURE classification with $k = 3$. In both analyses, individuals with African (AFR), European (EUR) and East Asian (EAS) ancestry the 1000 Genomes Project phase 3 (1KGP3) were used as parental references for ancestry inference. We considered only the overlapping SNPs included in the 654,027 Illumina's Infinium Global Screening Array across all cohorts for this analysis. For the PCA, the individual's scores in the first two principal components were plotted using R *ggplot2*.

### Linkage Disequilibrium (LD)

Linkage disequilibrium comparisons were performed between the three main continental ancestries 1KGP3 - African (AFR), European (EUR), East Asian (EAS) — and GRAR and SABE. We collected overlapping variants in all cohorts within a region of 500kbp upstream and downstream of the 313-SNPs used for the PRS and processed them with the varLD[25]. VarLD is a program for quantifying differences in LD patterns between populations. The steps involved in calculating the LD score include a) quantifying the LD (defined as the signed $r^2$ metric) between every pair of SNPs in each window for each pair of populations; b) producing a square matrix representing the correlation between the SNPs in each window for the respective population pair; c) performing an eigendecomposition of each matrix; d) measure the difference between the ranked eigenvalues of each matrix element and using it as an equality measure. This is the raw VarLD score for the SNP windows. The magnitude of this score can be interpreted as a measure of the dissimilarity between the correlation matrices. Therefore, the varLD scores can be interpreted as the extent of local LD differences between the populations. More details about raw varLD scores calculation are described elsewhere.

For plotting, we calculated the relative position of each neighbor variant to the focal risk SNP from the BC 313-SNP set, and used the *geom_smooth()* function from ggplot2 R package with a generalized additive model smoothing.

#### Regression model evaluation

To evaluate the PRS performance in the Brazilian and UKBB cohorts, we created regression models using the *glm()* function in R. Area under receiver operator curve (AUROC) was calculated using the *pROC* R package (1.18.0v). The 95% confidence intervals were calculated with the bootR package (1.3v). Briefly, we performed 1,000 bootstraps of samples from each cohort using the same number of cases and controls as the original cohorts. For AUROC and odds-ratio per standard deviation (ORperSD), a simple regression model with case status predicted by PRS only was chosen. For Partial-R2 two models were compared: a full model with *age + 5PCs + PRS* and a reduced model with *age + 5PCs* only. For PRS analysis samples from GRAR and SABE with East Asian ancestry fraction above 99%, inferred by ADMIXTURE [24], were removed, since our analysis was mainly focused on the admixed individuals.

#### Model comparisons and potential pitfalls

We described distributions of BC risk in models containing only the PRS, and PRS plus age and 5 PCs. We note that sampling methods among cohorts were significantly different and produced age distributions and a priori risks that might modulate risk distributions likely to be independent of the ancestry compositions themselves. SABE, although census-based, is focused on older individuals to which disease status is consolidated, particularly for phenotypes with adult to late onset such as BC. Therefore, SABE controls may reflect survival bias. Additionally, the restricted number of cases may affect the results due to low sampling power. In GRAR samples, cases and controls are younger, meaning that cases might be overrepresented by higher risk individuals, while part of the controls has a higher chance of manifesting the disease lately. To address potential confounders, we removed individuals with positive results for related cancers such as ovarian and pancreatic cancer and BC-cases with monogenic variants related to BC.

### Supplementary Tables

**Supplementary Table 1:** Demographic characteristics of the Brazilian cohorts

| Characteristic | **GRAR**, N = 853[1] | **SABE**, N = 753[1] | p-value [2] |
|---|---|---|---|
| Age (in years) | 40 (33, 50) | 72 (63, 80) | <0.001 |
| PRS | -0.11 (-0.51, 0.29) | -0.11 (-0.53, 0.34) | 0.7 |
| Breast Cancer | | | <0.001 |
| Case | 322 (38%) | 21 (2.8%) | |
| Control | 531 (62%) | 732 (97%) | |

[1] Median (IQR); n(%)

[2] Wilcoxon rank sum test; Pearson’s Chi-squared test

### Supplementary Figures

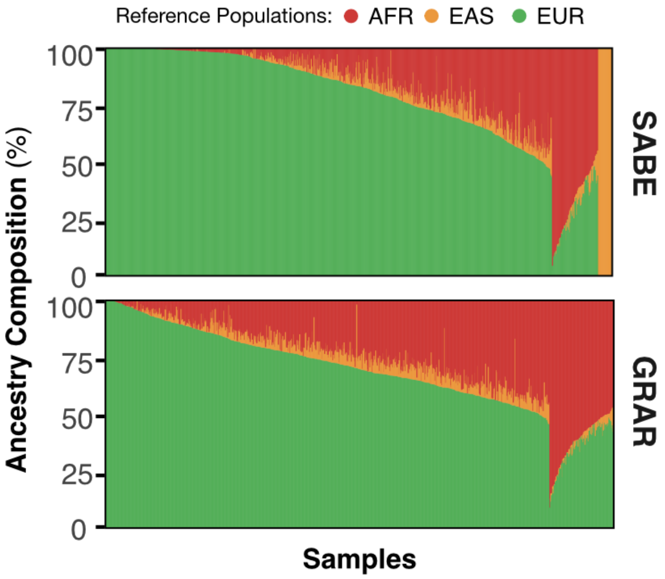


**Supplementary Figure 1: Global ancestry inference of GRAR and SABE cohorts.** Individual ancestry boxplots of SABE and GRAR cohort using supervised ADMIXTURE (k = 3) analysis shows that both SABE and GRAR individuals share similar patterns of ancestry proportions. Most individuals in both cohorts have a high proportion of European ancestry (above 75%) with a smaller fraction of African ancestry. SABE and GRAR has a distinct set of individuals with an almost exclusively East Asian ancestry component (orange cluster on top figure).

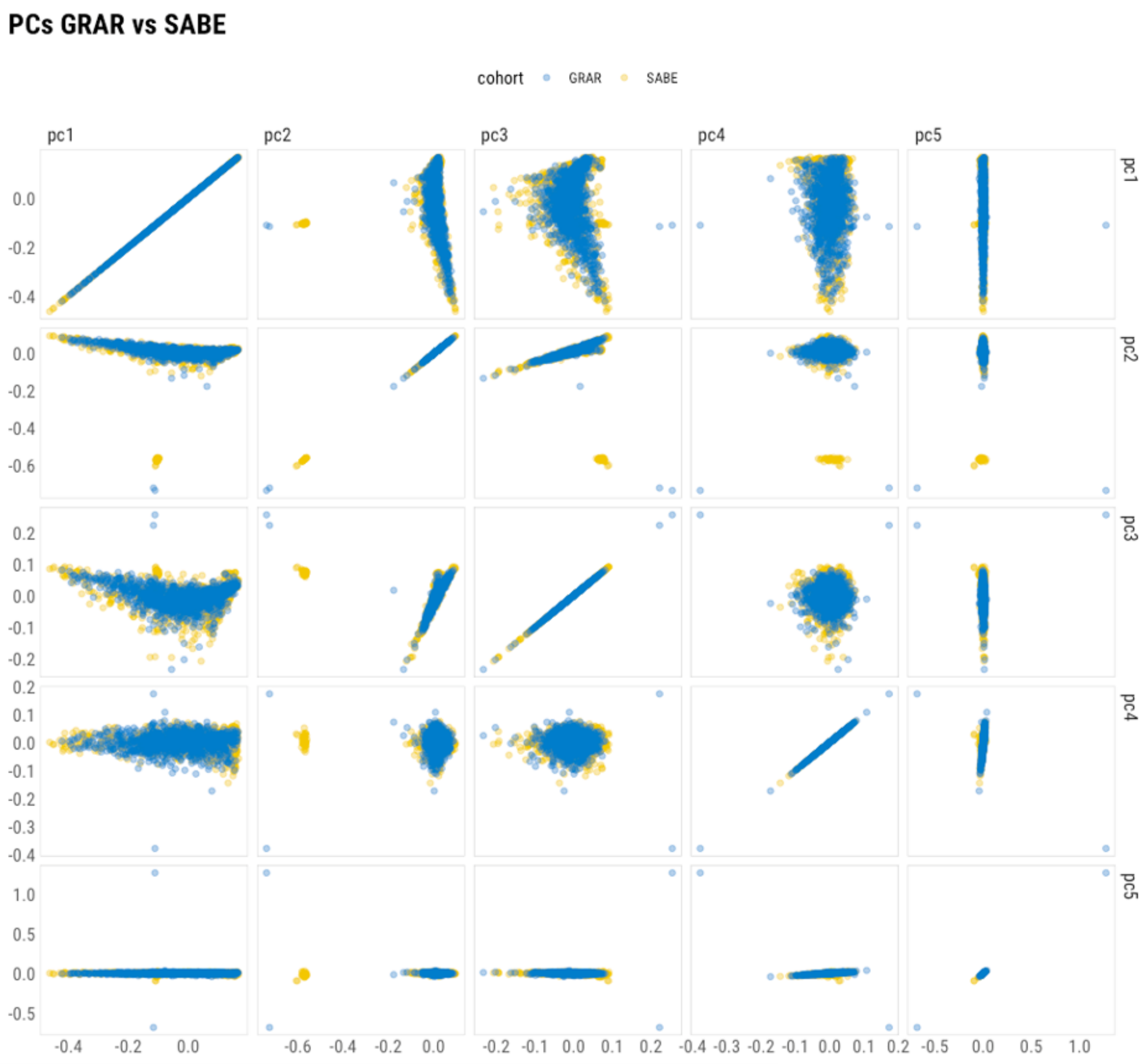


**Supplementary Figure 2: Genome-wide Principal Components on SABE and GRAR.** The panel shows the first five genome-wide principal components (PCs) for SABE (yellow) and GRAR (blue) individuals. The overlap position across the evaluated PCs between SABE and GRAR individuals evidences the similarity in ancestry composition between the two cohorts.

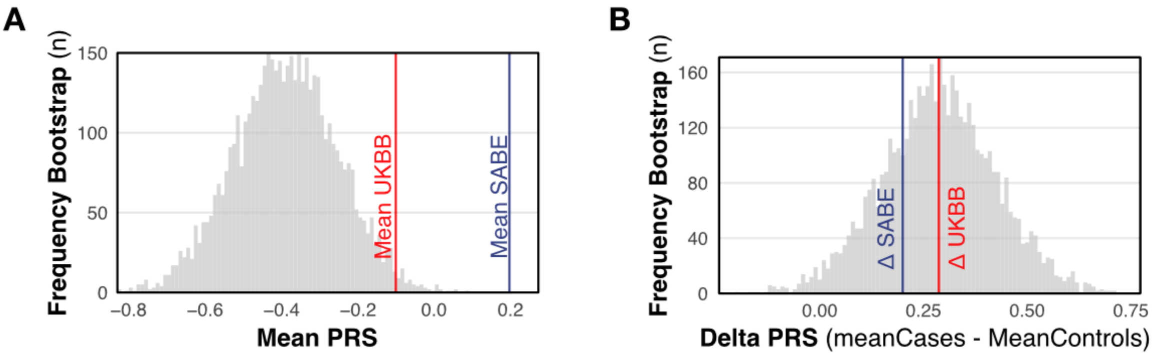


**Supplementary Figure 3: Bootstrap resamplings of the UKBB with the same total number and proportions of cases and controls as in SABE.** A) The distribution shows the mean 313-PRS values obtained in the 1,000 bootstrap resamplings B) The distribution shows the difference in mean 313-PRS between cases and controls (delta mean PRS) for 1,000 bootstrap resamplings. The red line represents the empirical values from the SABE cohort.

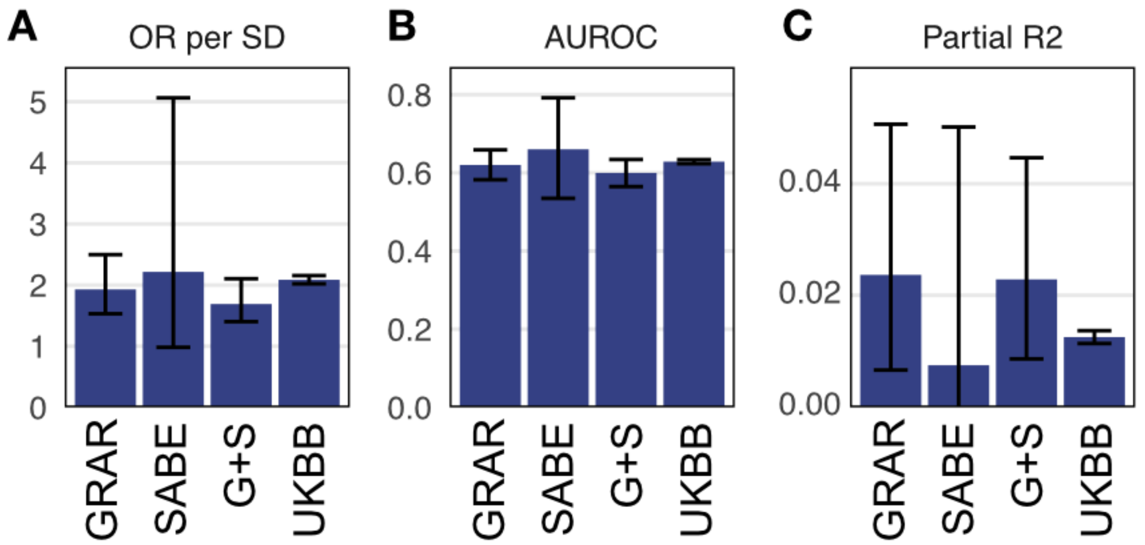


**Supplementary Figure 4: Predictive power of 313-PRS for all cohorts.** PRS showed a similar accuracy in all cohorts when considering the **(A)** odds-ratio per standard deviation and **(B)** AUROC, although the values reached a wider range in SABE with bootstrap analysis, as evidenced by the confidence interval. The partial-$R^2$ was calculated using a full model with case predicted by age + PRS + 5PCs and a reduced model with all variables but the PRS. The partial-$R^2$ was numerically higher in GRAR and G+S samples than in UKBB, unlike the observed in SABE **(C)**.
